# Cardiac MRI Phasic Assessment of Strain in Right Ventricular Dysfunction

**DOI:** 10.1101/2024.08.19.24312280

**Authors:** Alexandra M Janowski, Finley Mueller, Shreya Agarwal, Scott Visovatti, Rebecca R Vanderpool

## Abstract

**Background:** RV strain associates with mortality in pulmonary hypertension (PH) but time-resolved strain is not typically assessed. The aim was to evaluate phasic changes in RV strain using cardiac magnetic resonance (CMR) images. We hypothesized that phasic changes in ejection and filling RV strain significantly associate with outcomes in PH.

**Methods:** Participants were identified from the Ohio State University CMR PH registry (n=96). RV endocardial areas were segmented from 4-chamber CMR Cine images. Time-resolved strains were calculated for RV global, free wall and septal strain. Ventricular dynamics were assessed during the ejection, early filling and late filling cardiac phases to quantify phasic changes in function. RV contractility, afterload and diastolic stiffness were quantified using the single-beat method. Outcomes were evaluated at one year.

**Results:** In this retrospective, single-center study, 96 participants with and without pulmonary hypertension were included. Cohort was predominately female (n=53, 55%) with elevated mean pulmonary arterial pressure (38[26-48] mmHg) and reduced RV function (RVEF: 42[31-54] %, TAPSE of 19[15-23] cm). Filling strain patterns described changes in ventricular dynamics but did not associate with RV dilation or other measures characteristic of RV dysfunction. In comparison, decreased free wall strain and increased diastolic stiffness both associated with RV dysfunction but there were no significant differences in strain patterns. Participants with strain pattern 3, decreased free wall strain or increased Eed had increased one-year mortality. When investigated together, participants with decreased free wall strain, RVEF and increased Eed had greatly reduced one-year survival.

**Conclusions:** Assessment of phasic changes in ventricular function does provide additional pathophysiological information but assessment of strain patterns alone are not sufficient for identifying reduced function. Deep phenotyping using a combination of RV strain and diastolic stiffness is highly selective of participants with increased one-year mortality.

## Introduction

Right ventricular (RV) size and function associate with mortality and were included in the comprehensive risk assessment guidelines in the 2022/2023 Pulmonary Hypertension guidelines^1^ ^2,3^ The basic imaging and hemodynamic variables of stroke volume index and right atrial pressure help describe ventricular pump function and the relationship between cardiac output and preload pressure.^4,5^ However, advanced measures of RV systolic and diastolic function including RV strain and diastolic stiffness have the potential to improve pathophysiology-based assessment of right heart function and pump function. Increased diastolic stiffness has been shown to associate with RV dysfunction.^6–8^ Additionally, three post-systolic filling patterns have been identified that associate with the development of RV dysfunction with different patterns to distinguish between preserved RV function and end-stage right heart failure.^9^

Cardiac magnetic resonance (CMR) imaging is the gold standard for assessing RV size, shape, and function^3,10^ where changes in RV size and function throughout the cardiac cycle enables the phasic evaluation of ventricular dynamics.^11^ Assessment of RV strain from CMR images has been shown to have good agreement with echocardiographic RV free wall strain.^10,12,13^ When RV strain analysis was combined with single-beat analysis of RV diastolic function, Wessels et al. showed that increased RV diastolic stiffness associated with decreased right atrial function and alterations in RV filling.^11^ A recent study by Ghio S et al. demonstrated improvements in risk stratification models of PAH when pathophysiology-based approaches using advanced imaging were used.^14^ Comprehensive assessment of right ventricular function is improved with inclusion of both pressure and volume/structure data in RV functional assessments and endophenotyping patients with PH.

The overall aim of this study was to evaluate phasic changes in RV function with a focus on strain throughout the cardiac cycle via CMR images in patients with PH. The specific aims of this study were to 1) implemented RV strain analysis methods on four chamber CMR images. 2) identify phasic changes in RV function that associate with previously identified strain patterns^9^, free-wall strain and diastolic stiffness.

## Methods

### Study Population

In this retrospective cohort study, participants that underwent both CMR imaging and a right heart catheterization (RHC) at The Ohio State University (OSU) Medical Center were identified. (**Figure S1**). Individuals were first identified by searching clinical CMR report summary statements in the OSU CMR image database for keyword close variants of “pulmonary hypertension” between the years 2010-August 2021 (N = 235 participants). The OSU CMR image database included DICOM images, demographics, vital status, and structured data from clinical CMR reports including right and left ventricular volumes. Electronic records were reviewed for additional clinical data including right heart catheterization results and the clinical pulmonary hypertension diagnosis. Participants without right ventricular volume measurements were excluded (n = 74). The study was approved by The Ohio State University’s Institutional Review Board and informed consent was waived (IRB# 2021H0394).

### Four Chamber RV Endocardial Area Segmentation

CMR images were acquired as part of normal clinical care using Siemens 1.5-T scanners (Siemens Medical Solutions). Postprocessing on four-chamber cine CMR images was performed to identify the right ventricular endocardial area contours throughout the cardiac cycle using a custom MATLAB program (MATLAB R2021a, The MathWorks, Inc) (**Figure 1A**, blue overlay). The lateral and septal points of the tricuspid value were identified on the area contours (**Figure 1B**). The RV apex was used to separate the RV free wall (RV FW) from the septum. Segmented contours were used to determine RV area, RV global length (L _Total_), RVFW length (*L _FW_*), and septal length (*L _Sep_*) throughout the cardiac cycle (**Figure 1B**). Strain was calculated as the ratio of the change in length (L-L_0_) to the original length (L_0_) where L is the length at a given point in the cardiac cycle and L_0_ was set as the end-diastolic length. Strain was calculated for RV Global strain ([L_total_ – L_D total_]/L_D total_), RV free wall strain ([L_FW_ – L_D FW_]/L_D FW_) and RV septal strain ([L_Sep_ – L_D Sep_]/L_D Sep_) (**Figure 1B**).

**Figure 1.**
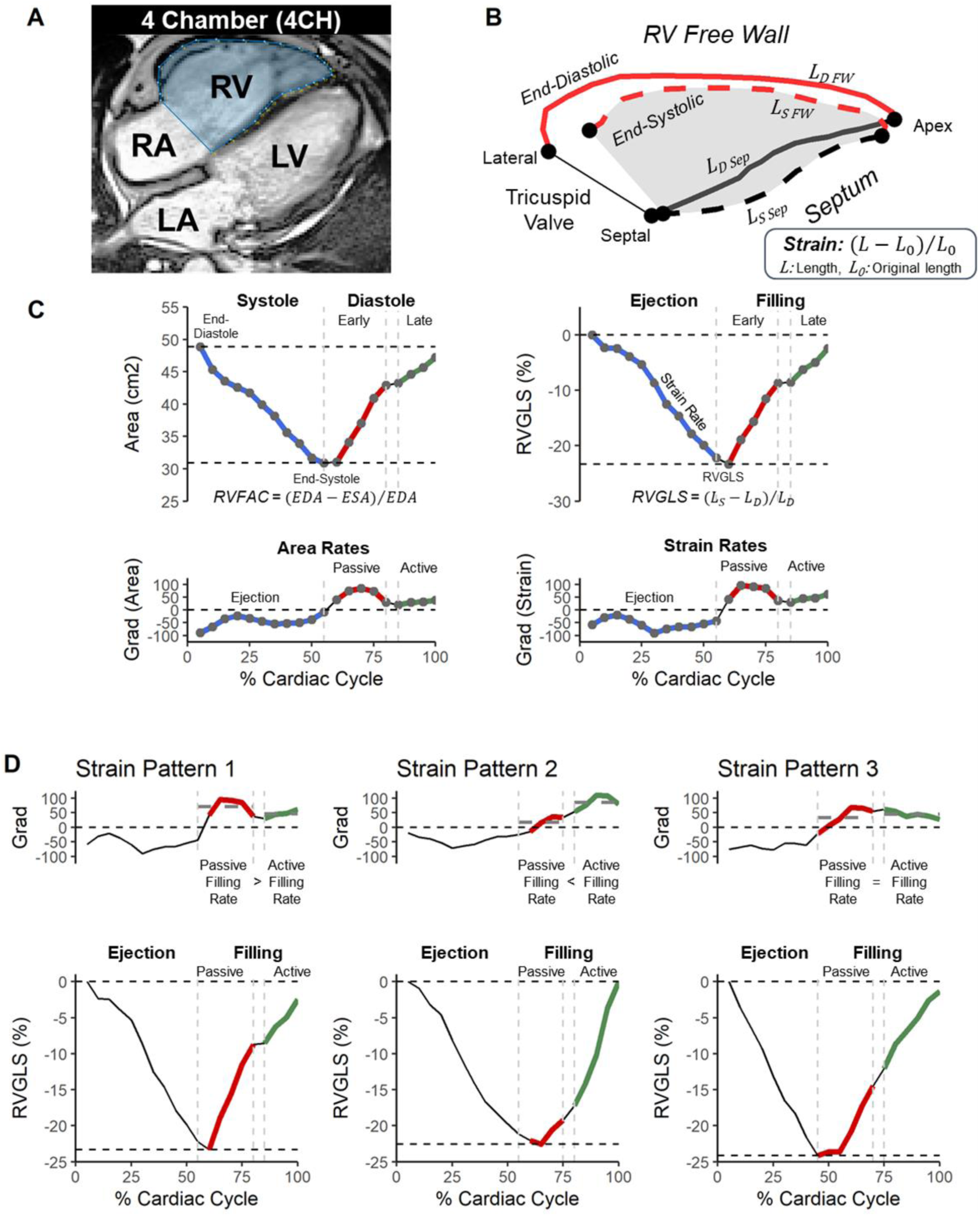
Analysis of RV area and strain from 4CH CMR images. A) RV endocardial area (blue area) was segmented from standard four chamber (4CH) CMR images in the complete cardiac cycle. **B)** Representative RV endocardial areas at end-diastolic and end-systolic with the lateral and septal tricuspid valve and RV apex landmarks identified. Total RV length was defined as the endocardial perimeter from the lateral to septal tricuspid valve points. RV free wall was the length from the lateral tricuspid valve to the RV apex and septal length was then the length from the RV apex to the septal tricuspid valve. Global, free wall and septal strain were calculated as the change in length minus the original length (end-diastole) divided by original length for the respective strain types. **C)** Area and Strain was assessed during the systolic (ejection, blue) and diastolic (filling, red and green) phases of the cardiac cycle. The diastolic filling phase was broken down into early/passive (Red) and late/active (green) phases. RV fractional area change (RVFAC) and RV global strain (RVGLS) was assessed at end-systole. The slope of the area (area rate) and strain curves were determined from the average gradient in each respective phase. **D)** The shape of the strain curve was then classified based on previously identified strain patterns^9^ based on the ratio of the passive and active strain rates. In strain pattern 1, passive filling strain rate is greater than active filling strain rate where strain pattern 2 is the opposite (passive < active). Strain pattern 3 has similar passive and active filling rates.

### RV Fractional Area Change - Ejection and Filling Area Rates

RV area curves were first normalized to percent cardiac cycle by interpolating to 5% cycle time increments using Piecewise Cubic Hermite Interpolating Polynomial (PCHIP) interpolation (**Figure 1C**). Normalization facilitated comparison between individuals and account for variable heart rates between participants. End-diastolic area (EDA) was defined as the largest RV area and end-systolic area (ESA) was defined as the smallest area. Fractional area change (FAC) was calculated as FAC = [EDA-ESA]/EDA. The time-resolved fractional area change (Grad - Area) was used to identify the ejection, passive filling and active filling phases (**Figure 1C**, area). The area ejection rate was calculated as the average of the negative derivatives during ejection (**Figure 1C**: linear decreasing area, blue). RV filling rates were split into passive filling rate (red) and active filling rate (green). The passive filling rate was calculated as the average of the positive derivatives during early diastole and the active filling rate was calculated as the average of the positive derivatives during late diastole.

### Maximum Strain and Strain Rates

Maximum strain values are reported at end-systole where strain is the difference in length at end-diastole and end-systole for global strain (RV Global Strain), free wall strain (RVFW Strain) and septal strain (RV Septum Strain) (**Figure 1C**, RVGLS example). Strain rates were calculated as the rate of change in strain over the ejection, passive filling, and active filling phases. From the time-resolved strain rate, ejection strain rate was calculated as the average of the negative derivatives during ejection (**Figure 1C**: blue). Passive filling strain rate was calculated as the average of the positive derivatives during early diastole (red) and active filling strain rate was calculated as the average of the positive derivatives during late diastole (green).

### Post-Systolic Strain Curve Morphology

Time-resolved strain curves were classified into three previously identified post-systolic strain patterns^9^ (**Figure 1D**). Strain pattern 1 is characterized by a steep passive filling strain rate and transitions to a shallow active filling strain rate (passive filling rate > active filling rate). Strain pattern 2 is characterized by the opposite, a shallow passive filling strain rate and a steep active filling strain rate (passive filling rate < active filling rate). Strain pattern 3 is characterized by linear filling with equal passive and active filling strain rates (passive filling rate = active filling rate). In terms of clinical relevance, strain pattern 1 associates with normal RV size and function, strain pattern 2 associates with severe PH with preserved filling pressure and cardiac index and strain pattern 3 associates with increased mortality, RV dilation, and reduced RV systolic function.^9^

### Hemodynamic Assessment

Right heart catheterization reports were reviewed to obtain pulmonary arterial pressure (PAP), pulmonary arterial wedge pressure (PAWP), RA pressure, RV pressure, and thermodilution and Fick cardiac outputs. Thermodilution cardiac output measurements were used but Fick cardiac outputs were used when thermodilution cardiac output was not available. Stroke volume (SV) was calculated as cardiac output divided by heart rate. Pulmonary vascular resistance (PVR) was calculated as the difference between mean PAP and PAWP divided by CO. Pulse pressure was defined as the difference between systolic and diastolic PAP. Pulmonary arterial compliance (PACa) was calculated as the stroke volume divided by the pulse pressure. Cardiac index was defined as CO divided by BSA. Participants were grouped based on the 2022 ERS/ESC guidelines hemodynamic definitions of pulmonary hypertension with a mPAP cut-off of 20 mmHg (No PH: mPAP ≤ 20mmHg).^1,15^ Participants with pre-capillary PH hemodynamics had elevated mPAP, normal PAWP and elevated PVR (mPAP > 20mmHg, PAWP ≤ 15 mmHg, PVR > 2WU). Participants with PH and elevated Pulmonary artery wedge pressure (PAWP > 15mmHg) were split into isolated post-capillary PH (ipc-PH: PVR ≤ 2WU) and combined pre/post-capillary PH (cpc-PH: PVR > 2WU). Participants that did not meet these hemodynamic definitions (mPAP > 20mmHg, PAWP < 15mmHg, PVR ≤ 2WU were combined with the ipc-PH group.

### Single-beat assessment of RV function

RV pressure waveforms were obtained and digitized using Matlab to quantify RV function using the single beat method.^6,7,16,17^ Max isovolumic pressure (Pmax) was found by fitting a sinusoidal curve to the RV waveform during early isovolumic contraction and late isovolumic relaxation phases. End-systolic pressure (ESP) was estimated from mean PA pressure with 1.65 (mPAP) - 7.79.^18^ End systolic elastance (Ees), a measure of RV contractility, was calculated as the ratio of (Pmax-ESP) to SV. Arterial elastance (Ea), a measure of RV afterload, was calculated as ESP/SV. The RV-PA coupling ratio (Ees/Ea) was calculated as (Pmax-ESP)/ESP. RV diastolic stiffness (β) was determined by fitting a non-linear exponential curve (P=α(e^Vβ^-1)) where α is a curve-fitting parameter and β is the diastolic stiffness coefficient^6,7,19^. End-diastolic elastance (Eed) was calculated as the slope of the end-diastolic pressure-volume curve, P=αβ(e^Vβ^), using right atrial pressure as an estimate for RV end-diastolic pressure and CMR end-diastolic and end-systolic volumes.^17^

### Statistical analysis

Continuous data is presented as median [IQR], and categorical variables are presented as the number of participants (percentage). Significance was measured via Kruskal Wallis and Dunn tests. Fisher’s exact test was used to assess significance for categorical variables in R. A p-value of 0.05 was considered significant. To compare RV function characteristics the strain pattern groups, participants were grouped by RVFW strain (FW-G1, FW-G2, and FW-G3) and Eed groups (Eed-G1, Eed-G2 and Eed-G3). Concordance between strain patterns, RV FW strain and Eed in classifying participants was investigated using an Alluvial plot.

Follow-up was assessed as time between the index CMR imaging and death or last day of follow-up (May, 2024). Outcomes analysis focused on one-year survival. Univariable Cox proportional hazards regression was performed to identify variables that associate with one-year mortality. Multi-variable Cox Proportional Hazards regression was performed on variables that significantly associated with mortality with hazard ratio p-value of < 0.05. Kaplan Meier analysis with pairwise Log-rank tests were performed to evaluate differences in mortality between groups.

## Results

### Cohort Characteristics

A total of 96 participants with 4-chamber CMR images and RHC data were included (**Table 1**). Participants were excluded if they were missing RV or LV volumes, no DICOM images, no RHC data or had poor signal quality (N = 129) (**Figure S1B**). The final cohort was predominately female (n=53, 55%) with elevated mean pulmonary arterial pressure (38 [26-48] mmHg) (**Table 1**). Overall, RV function is reduced with a median RV ejection fraction of 42 [31-54] %, TAPSE of 19 [15-23] cm, and RV-PA coupling of 0.72[0.4-1.3].

**Table 1.** Cohort Characteristics across Strain Patterns.

|  | **Overall**<br>**N = 96** | **Pattern 1**<br>**N = 28** | **Pattern 2**<br>**N = 32** | **Pattern 3**<br>**N = 36** |
| --- | --- | --- | --- | --- |
| Age (years) | 56 [46, 67] | 55 [44, 67] | 59 [50, 67] | 55 [46, 66] |
| Female (%) | 66 (68.8) | 18 (64.3) | 23 (71.9) | 25 (69.4) |
| Heart rate (bpm) | 81 [71, 90] | 78 [65, 84] | 82 [73, 91] | 82 [73, 96] |
| BMI (kg/m<sup>2</sup>) | 28 [24, 33] | 26 [23, 32] | 28 [26, 33] | 31 [24, 35] |
| Survival at 1 year (%) | 80 (83.3) | 26 (92.9) | 29 (90.6) | 25 (69.4) |
| RA pressure (mmHg) | 8 [5, 14] | 8 [4, 11] | 8 [5, 13] | 11 [5, 18] |
| Mean PAP (mmHg) | 38 [26, 48] | 30 [20, 42] | 40 [35, 53]* | 38 [25, 49] |
| PAWP (mmHg) | 12 [9, 17] | 10 [6, 13] | 14 [10, 19]* | 13 [10, 21]* |
| Cardiac Index (L/min/m<sup>2</sup>) | 2.65 [2.01, 3.28] | 2.46 [2.04, 3.00] | 2.83 [2.08, 3.41] | 2.63 [1.87, 3.30] |
| PVRi | 8.3 [4.9, 14.7] | 8.3 [2.6, 18.0] | 9.1 [6.2, 14.4] | 7.7 [3.5, 12.3] |
| TAPSE | 18.7 [14.9, 22.4] | 21.0 [16.3, 26.5] | 18.8 [15.3, 22.4] | 17.3 [13.8, 21.0] |
| TAPSE/sPAP | 0.30 [0.20, 0.52] | 0.45 [0.24, 0.87] | 0.28 [0.19, 0.38] | 0.29 [0.20, 0.54] |
| RVEF | 42 [31, 54] | 42 [31, 55] | 42 [31, 53] | 42 [31, 52] |
| RVESVi | 57 [35, 74] | 57 [39, 67] | 56 [36, 72] | 57 [31, 76] |
| SVI | 38 [31, 46] | 39 [32, 48] | 39 [31, 49] | 36 [30, 41] |
| SV/ESV | 0.74 [0.46, 1.19] | 0.73 [0.46, 1.25] | 0.74 [0.45, 1.11] | 0.72 [0.46, 1.08] |
| RV Global Strain/sPAP | 0.34 [0.22, 0.58] | 0.45 [0.27, 0.97] | 0.31 [0.20, 0.49]* | 0.35 [0.22, 0.63] |
| Pmax | 112 [83, 160] | 98 [55, 130] | 132 [102, 179] | 102 [83, 164] |
| ESP | 55 [35, 71] | 43 [25, 62] | 58 [50, 80]* | 56 [34, 73] |
| Ees | 0.73 [0.41, 1.33] | 0.52 [0.29, 1.25] | 0.86 [0.51, 1.27] | 0.75 [0.46, 1.46] |
| Ea | 0.71 [0.42, 1.10] | 0.56 [0.25, 0.98] | 0.82 [0.55, 1.19] | 0.69 [0.45, 1.01] |
| Ees/Ea | 1.03 [0.69, 1.53] | 1.38 [0.74, 1.97] | 1.04 [0.58, 1.52] | 0.99 [0.73, 1.38] |
| Eed | 0.27 [0.08, 0.47] | 0.19 [0.08, 0.39] | 0.25 [0.09, 0.46] | 0.31 [0.09, 0.75] |

BSA, body surface area; BMI, body mass index; RA, right atrium; PAP, pulmonary artery pressure; PAWP, pulmonary artery wedge pressure; PVRi, pulmonary vascular resistance index; TAPSE, tricuspid annular plane systolic excursion; EF, ejection fraction; ESVi, end systolic volume index; SVI, stroke volume index; Pmax, max isovolumetric pressure; ESP, end-systolic pressure; Ea, arterial elastance; Ees/Ea, RV-PA coupling ratio; Eed, end diastolic elastance. \*p < 0.05 vs Strain pattern 1, †p < 0.05 vs Strain pattern 2,

### Phasic Right Ventricular Function – Strain Patterns and Free-Wall Strain

When classified by the post-systolic strain patterns, all three stain patterns were present but more participants classified as having strain pattern 3 (**Table 1**, n = 38 participants). RV end-diastolic area, end-systolic area and RV fractional area change were not significantly different between strain patterns (**Figure 2A**, **Table 1**). Additionally, there were no significant differences in RV global strain (**Figure 2B**, p = 0.16). Phasic changes in RV function including ejection, passive filling and active filling strain rates were explored in **Figure 2C** and **Table 2**. Within each strain pattern, the relationship between passive and active strain are consistent with the selection criteria (**Figure 2C)** where pattern 1 has greater passive than active filling strain rate, pattern 2 has a greater active than passive filling strain rate, and pattern 3 has similar passive and active filling strain rates (**Figure S2A)**. Comparing across strain patterns, passive filling strain rate is increased in strain patterns 2 and 3 compared to strain pattern 1 (**Figure 2C**). For the active filling strain rate, strain pattern 2 had the highest rate with no significant differences between strain patterns 1 and 3. Strain patterns 2 and 3 have decreased ejection strain rates compared to strain pattern 1 (**Figure 2C**, **Table 2**, p = 0.003 and p = 0.02, respectively). For other clinical characteristics, there were no significant differences in age, sex, and BMI between strain patterns (**Table 1**). Additionally, there were no significant differences in PVRi between strain patterns despite elevated mean PA pressure (Pattern 2) and elevated PAWP (Patterns 2 and 3) in some of strain patterns. Similarly, there were no significant differences in RV contractility (Ees), arterial afterload (Ea), RV-PA coupling (Ees/Ea) or diastolic stiffness (Eed) between strain patterns. Post-systolic filling strain patterns describe changes in ventricular dynamics but did not associate with RV dilation or other measures characteristic of RV dysfunction.

**Figure 2.**
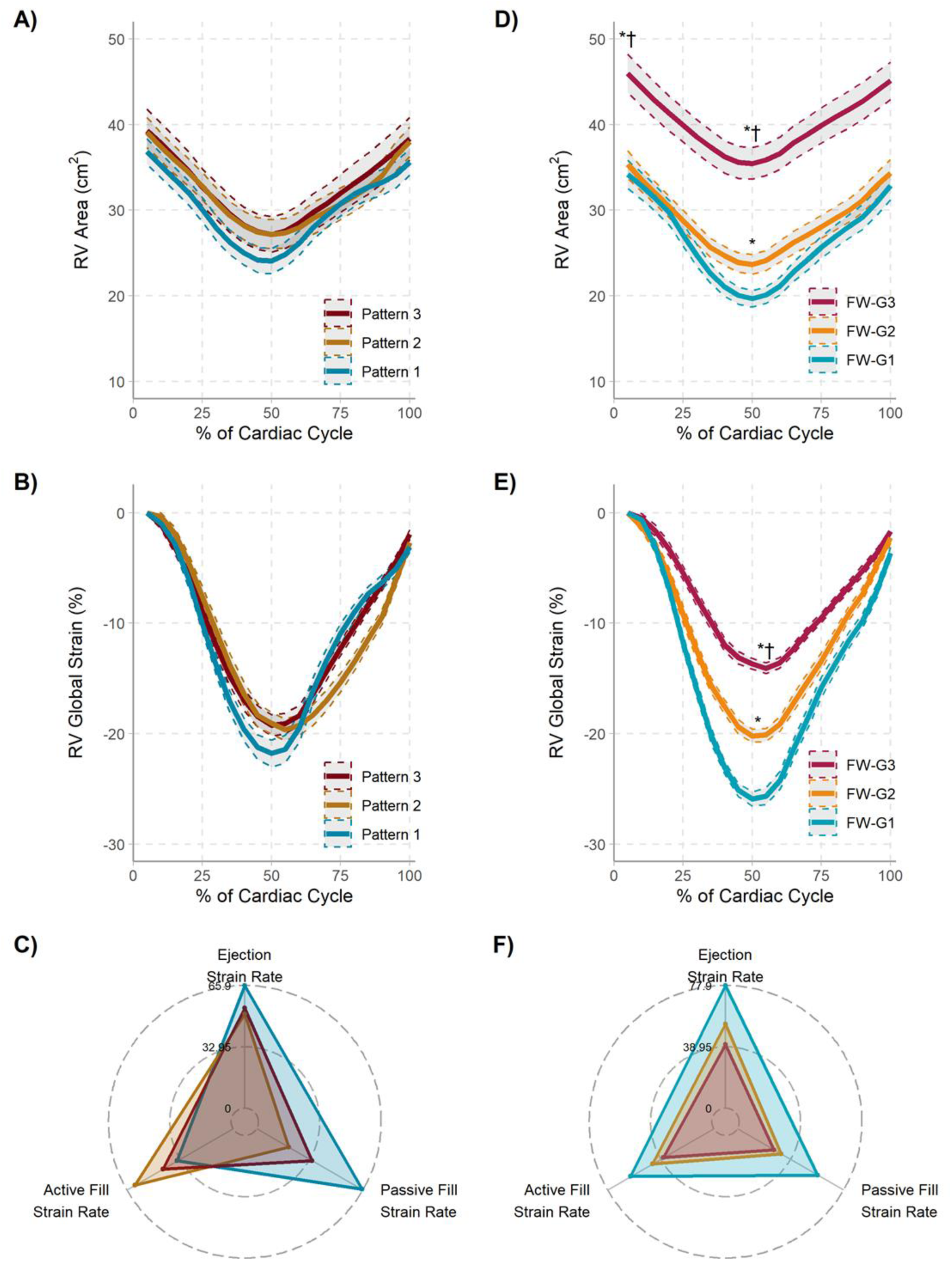
Characterization of phasic right ventricular function in strain pattern and RV free wall strain groups. A) When split by strain patterns, there were no significant differences in end-diastolic or end-systolic area. Pattern 1: blue, Pattern 2: yellow, Pattern 3: Red. **B)** RV global strain curves demonstrate the differences in filling patterns for the strain patterns. Pattern 1 has a steep passive filling strain rate with a shallow active filling strain rate. Pattern 2 has a shallow passive filling strain rate with a steep active filling strain rate. Passive and active filling strain rates are similar in Pattern 3. Despite differences in the selected filling patterns, there were no significant differences in max strain. **C)** Phasic differences in ejection/filling strain rates were further explored in spider plots. Passive and active filling strain rates reflect the selection criteria with a high passive filling rate in pattern 1 and high active filling rate in pattern 2. In pattern 3, there were not significant differences in passive and active filling strain rates. When compared across strain patterns, ejection strain rate was significant decreased in Patterns 2 and 3 compared to Pattern 1. Statistical comparisons are further explored in Table 2. **D)** When classified by RV FW strain, participants with decreased strain have increased RV areas. FW-G1: blue, FW-G2: yellow, FW-G3: Red. *p<0.05 v FW-G1; †p<0.05 v FW-G2. **E)** RV global end systolic strain is significantly decreased in FW-G2 and further decreased in FW-G3 compared to FW-G1. *p<0.05 v FW-G1; †p<0.05 v FW-G2. **F)** Within each FW group, there were no significant differences in passive and active filling strain rates. Ejection, Passive filling and Active filling strain rates are significantly decreased in FW-G2 and 3 compared to FW-G1. Statistical comparisons are further explored in Table 2.

**Table 2.** Strain and Strain Rates between Strain Patterns, Free Wall (FW) strain groups and Diastolic Stiffness (Eed) groups.

|  | n | RV Global Strain | RV FW Strain | Ejection Strain Rate | Passive Filling Strain Rate | Active Filling Strain Rate |
| --- | --- | --- | --- | --- | --- | --- |
| Overall | 96 | 21 [16, 25] | 22 [18, 29] | 52 [41, 70] | 32 [19, 52] | 40 [30, 57] |
| Pattern 1 | 28 | 22 [18, 27] | 27 [19, 33] | 71 [49, 88] | 61 [48, 80] | 34 [19, 48]<sup>#</sup> |
| Pattern 2 | 32 | 21 [16, 25] | 22 [18, 26] | 44 [38, 59]<sup>*</sup> | 19 [10, 26]<sup>*</sup> | 56 [40, 72]<sup>*#</sup> |
| Pattern 3 | 36 | 21 [15, 24] | 22 [15, 28] | 52 [38, 66]<sup>*</sup> | 32 [23, 43]<sup>*†</sup> | 38 [32, 52]<sup>†</sup> |
| FW-G1 | 32 | 27 [25, 30] | 32 [29, 37] | 74 [63, 92] | 56 [34, 74] | 55 [37, 75] |
| FW-G2 | 32 | 21 [19, 22]<sup>*</sup> | 22 [21, 25] | 52 [44, 61]<sup>*</sup> | 31 [19, 40]<sup>*</sup> | 40 [30, 59] |
| FW-G3 | 32 | 14 [13, 16]<sup>*†</sup> | 15 [12, 18] | 37 [28, 46]<sup>*†</sup> | 24 [18, 36]<sup>*</sup> | 35 [26, 40]<sup>*</sup> |
| Eed-G1 | 32 | 23 [19, 27] | 26 [21, 30] | 60 [49, 74] | 31 [22, 50] | 38 [27, 61] |
| Eed-G2 | 32 | 23 [17, 26] | 25 [18, 32] | 55 [44, 77] | 38 [25, 69] | 49 [35, 64] |
| Eed-G3 | 32 | 16 [14, 20]<sup>*†</sup> | 18 [14, 22]<sup>*†</sup> | 41 [29, 58]<sup>*†</sup> | 24 [18, 44]<sup>†</sup> | 35 [26, 46]<sup>†</sup> |
| No PH | 14 | 28 [25, 31] | 35 [30, 38] | 85 [72, 94] | 60 [40, 78] | 42 [30, 59] |
| ipc-PH and other | 8 | 24 [19, 28] | 29 [21, 34] | 78 [56, 92] | 47 [30, 78] | 44 [34, 57] |
| cpc-PH | 24 | 18 [15, 22]<sup>*†</sup> | 20 [15, 24]<sup>*†</sup> | 48 [35, 61]<sup>*†</sup> | 25 [17, 38]<sup>*†</sup> | 45 [33, 56] |
| Pre-capillary | 50 | 20 [16, 24]<sup>*</sup> | 21 [17, 26]<sup>*</sup> | 49 [38, 62]<sup>*†</sup> | 30 [20, 44]<sup>*</sup> | 38 [26, 57] |

FW: Free wall strain groups, Eed: End-diastolic elastance groups.
For strain Patterns: \*p < 0.05 vs Strain pattern 1, †p < 0.05 vs Strain pattern 2.
For FW strain groups: \*p < 0.05 vs FW-G1, †p < 0.05 vs FW-G 2.
For Eed groups: \*p < 0.05 vs Eed-G1, †p < 0.05 vs Eed-G2,
For PH types: \*p < 0.05 vs No PH, †p < 0.05 vs ipc-PH and other, ‡ p < 0.05 vs cpc-PH ,
Within individual groups: #active filling strain rate p < 0.05 vs passive filling strain rate

In comparison, when participants were split by free-wall strain (FW-G1: 32 [30-37]%, FW-G2: 22[21-25]% and FW-G3: 15 [12-18]%), a decrease in free wall movement/strain (FW-G3) associated with an increase in end-diastolic area (FW-G2 and FW-G1, p<0.05) (**Figure 2D**). Decreased free wall movement (FW-G3) also associated with decreased RVFAC and RVEF (**Table S2**). Because free wall strain and global strain are highly correlated (Pearson r = 0.95), RV global strain was significantly decreased in FW-G2, and further decreased in FW-G3 compared to FW-G1 (**Figure 2E**). Within each RV free wall strain group, there were no significant differences between passive and active filling strain rates (**Table 2** and **Figure S2B**). When compared across the RV free wall groups (**Figure 2F**), passive and active filling stain rates are decreased in FW-G2 and FW-G3. RV ejection strain rate is decreased in FW-G2, with a further decrease in FW-G3 compared to FW-G1. There were no significant differences in age, sex, and BMI between free wall strain groups (**Table S2**). In contrast to strain patterns, PVRi significantly increased with decreased RV free wall strain (p<0.05). Contributing to the increased PVRi, mean PA pressure was significantly increased with decreased RV strain but with no significant differences in PAWP between FW strain groups. While there were no significant differences in RV contractility (Ees), arterial elastance (Ea) as significantly increased resulting in a significant decrease in RV-PA coupling in FW-G3 (0.71 [0.33-1.35] compared to FW-G1 (**Table S2**). End-diastolic elastance increased with decreased strain in FW-G2 and FW-G3 (p<0.05) compared to FW-G1. While decreased free wall strain associated with more RV dysfunction (decreased RVEF, RV FAC, increased PVRi and Eed), there were no significant differences in the post-systolic filling patterns that were present with strain patterns.

### Phasic Right Ventricular Function – Diastolic Stiffness

In the diastolic stiffness groups (Eed-G1 (0.05[0.02-0.08]), Eed-G2 (0.27[0.19-0.23]), and Eed-G3 (0.67[0.48-0.96]), RV end-diastolic and systolic area associated with increased diastolic stiffness (Eed-G2 and Eed-G3 vs Eed-G1, p<0.05) (**Figure 3A**). Related to the increased end-diastolic area, RV fractional area change (RVFAC) was significantly reduced in Eed-G3. When the full RV volume was considered, RVEF was also decreased in Eed-G2 compared to Eed-G1. When looking at strain, RV global strain was decreased in Eed-G3 compared to Eed-G1 and Eed-G2 (**Figure 3B and Table 2**) but there were no significant differences in phasic function. Within each Eed group, there were no significant differences between passive and active filling strain rates (**Figure 3C and Figure S2C**). When compared across Eed groups, RV ejection, passive filling and active filling strain rates are significantly decreased in Eed-G3 with no significant differences between Eed-G2 and Eed-G1 (**Figure 3C**, **Table 2**). There were no significant differences in age and sex between Eed groups except Eed-G3 had increased BMI. (**Table S2**). Similar to FW groups, increased PVRi associated with increased Eed with the highest PVRi in Eed-G3 (12.3 [9.4-18.2] WU*m^2^). Both mean PA pressure and PAWP were increased in Eed-G2 and Eed-G3 compared to FW-G1. While there were no significant changes in RV contractility (Ees), arterial elastance (Ea) was significantly increased in Eed-G3 (0.71 [0.46-1.04]) that resulted in a significant decrease in RV-PA coupling compared to Eed-G1 (**Table S2**).

**Figure 3.**
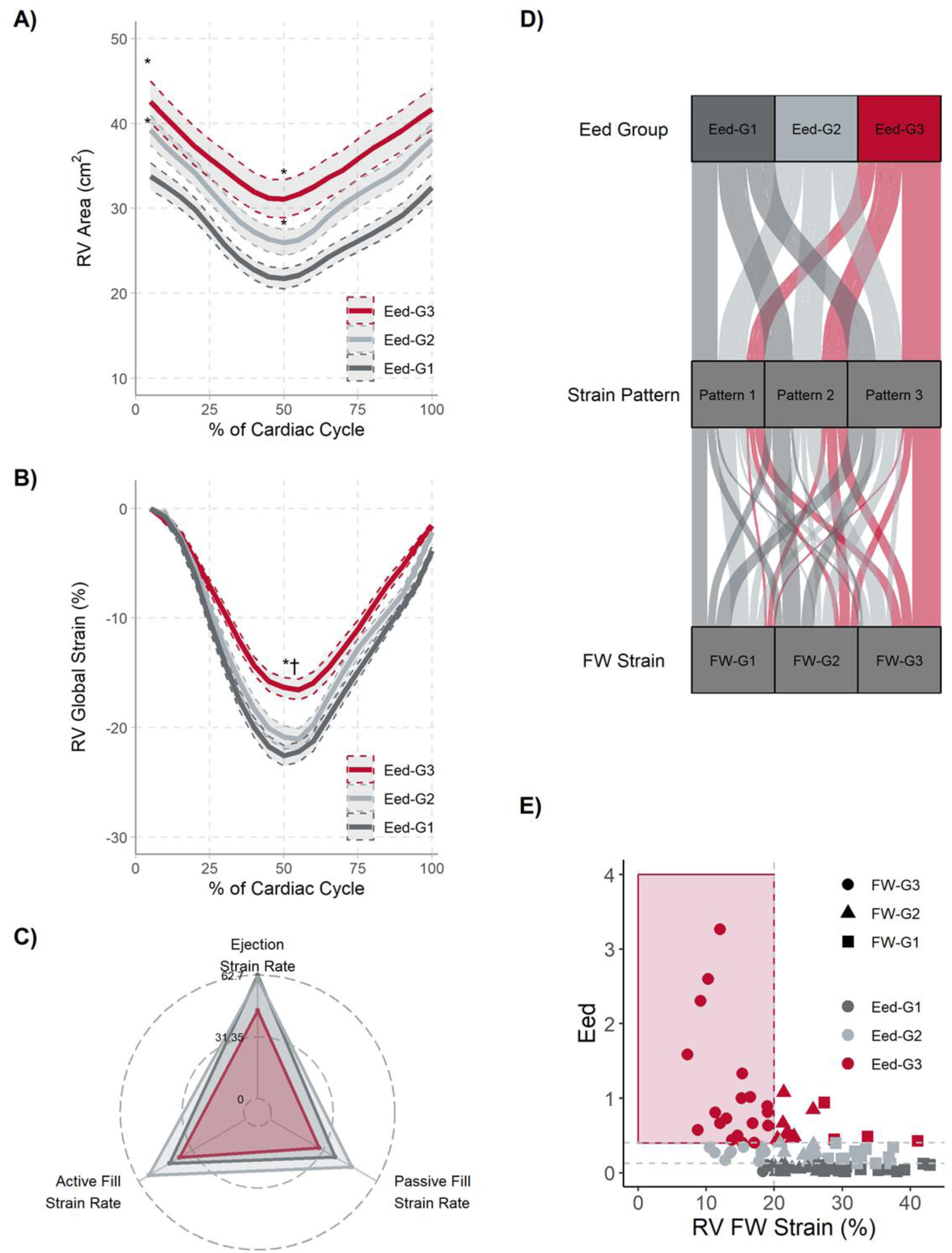
Association of diastolic stiffness with phasic RV function and strain. A) When split by diastolic stiffness (Eed), RV end-diastolic and end-systolic area are increase in Eed-G2 (light gray) and Eed-G3 (scarlet) compared to Eed-G1 (dark gray). **B)** For RV global end-systolic strain, RVGLS was significantly decrased in Eed-G3 compared to Eed-G1. There were no significant differences between Eed-G2 and Eed-G1. *p<0.05 compared to Eed-G1, †p<0.05 compared to Eed-G2. **C)** Within each Eed group, there were no significant differences in passive and active filling strain rates. Ejection, Passive filling and Active filling strain rates are significantly decreased in Eed-G3 compared to FW-G2. Statistical comparisons are further explored in Table 2. **D)** The concordance between diastolic stiffness, strain patterns and FW strain were investigated. Each strain pattern includes on average a third of the participants from each Eed group. Similarly, each FW group includes on average a third of the participants from each strain pattern but not always in concordance with the Eed-Groups. FW group 1 is mostly composed of participants with low diastolic stiffness (Eed-G1 and Eed-G2). FW-G3 has a higher percentage of participants with increased diastolic stiffness (Eed-G3). **E**) Eed significantly correlates with RV free wall strain (r = -0.48, p < 0.001).

### Intersection of Diastolic Stiffness, Post-Systolic Strain Patterns and Free Wall Strain on Phasic RV function

To better understand the intersection of diastolic stiffness and strain, the concordance of RV function metrics in individual participants were investigated (**Figure 3D**). On average, a third of the participants in each Eed group were classified into each of the strain patterns resulting in no significant differences in Eed between strain patterns. Similarly, a third of the participants in each FW group were classified into each of the strain patterns resulting in no differences in FW strain between strain patterns. Eed is negatively correlated with RV FW strain (r = -0.48, p < 0.001) where Eed increases at lower strains (**Figure 3E**). Participants in FW-G1 have low diastolic stiffness (0.12 [0.04-0.27] mmHg/ml) with participants mostly classified in Eed-G1 and Eed-G2 (87.5%). While there were more participants from Eed-G3 (increased from 12.5% to 28.1%), Eed was not significantly different in FW-G2 (0.18 [0.07-0.43] mmHg/ml) compared to FW-G1. FW-G3 is mostly composed of participants in Eed-G3 (59.4%) resulting in an increase in diastolic stiffness (0.47 [0.29-0.83] mmHg/ml).

To further investigate the interaction between ventricular dynamics and diastolic stiffness, we split the cohort by strain patterns and Eed groups (**Figure 4**). In strain patterns 1 and 2, there were no significant changes in RV area (**Figure 4A**) or RV global strain (**Figure 4B**) across Eed groups. In strain pattern 3, Eed-G3 had increased end-diastolic and end-systolic area compared to Eed-G1 and decreased global strain compared to Eed-G1 and Eed-G2 Due to the correlation between free wall strain and Eed, there were no significant changes in area or strain between Eed groups in each FW group (**Figure S3**). When investigated as a function of free-wall strain, there were no significant differences in end-diastolic or end-systolic area between Eed groups (**Figure S3A**). Overall, the strain curve morphology and global strain values remain consistent across Eed groups in each FW strain group. Because strain patterns do not associate with free wall strain or diastolic stiffness, strain patterns can be used to further classify differences in ventricular dynamics in participants.

**Figure 4.**
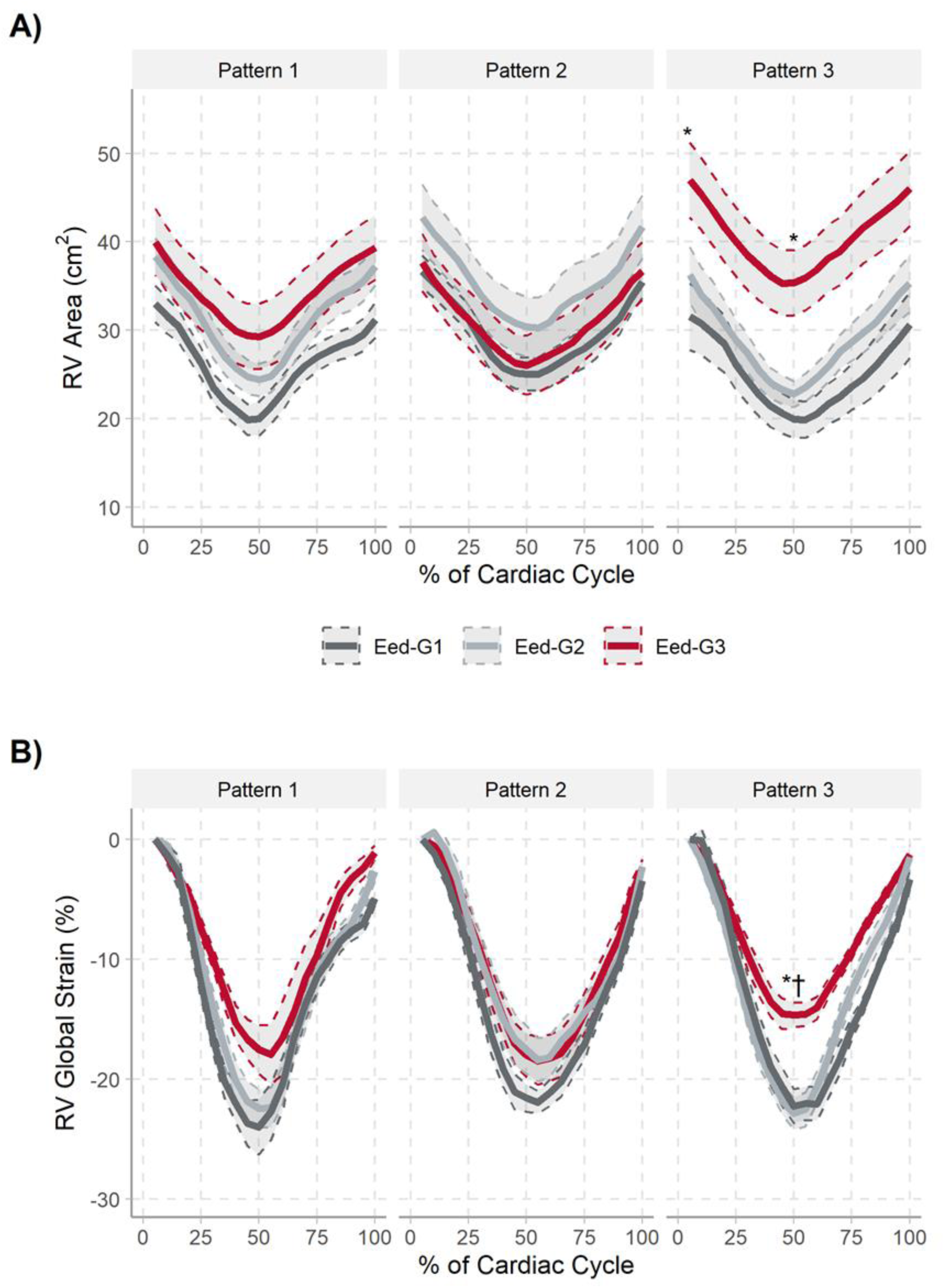
Intersection of strain patterns and diastolic stiffness on RV area and global strain A) In strain patterns 1 (Pattern 1) and 2 (Pattern 2), there were no significant differences in RV area between diastolic stiffness groups (Eed-G1: dark gray, Eed-G2; light gray, or Eed-G3, scarlet). RV end-diastolic and end-systolic area are increased in Eed-G3 in strain pattern 3 (Pattern 3). **B)** No significant differences in RV global end-systolic strain between Eed groups in strain patterns 1 and 2. In strain pattern 3, RV global strain is decreased in Eed-G3 compared to Eed-G1 and Eed-G2. *p<0.05 compared to Eed-G1, †p<0.05 compared to Eed-G2

### Association of Strain and Diastolic Stiffness with One-Year Mortality

Overall follow-up was 2.3 [0.97-5.6] years with 43 deaths at one-year follow-up there were 16 deaths. As continuous variables, RV global strain, Free wall strain and Eed are also associated with one-year mortality. In terms of ventricular dynamics, ejection strain rate was significantly associated with mortality, but passive and active filling strain rates did not. Using univariable Cox regression analysis, strain pattern 3, FW group 3, and Eed group 3 significantly associate with one-year mortality (**Table 3**). Kaplan Meier analyses show that strain patterns (**Figure 5A**), decreased FW strain (**Figure 5B**) and increased diastolic stiffness (**Figure 5C**) significantly associated with one-year mortality. When considering the intersection of strain and diastolic stiffness, participants with reduced strain and increased stiffness (FW strain < 20, Eed >0.4) have increased one-year mortality (**Figure 5D**). Multiple measures of RV function significantly associate with mortality with c-indices > 0.67 making it difficult to select individual or groups of variables. From the imaging variables recently included in the 2022/2023 Pulmonary Hypertension guidelines, TAPSE/sPAP, RVEF, RV ESVI and RA pressure significantly associate with one-year mortality. However, there was little concordance between the variables in individual participants (**Figure S4A**).

**Table 3:** Cox proportional hazards ratios for one-year survival for RV function variables and across strain patterns, Eed groups, and FW groups.

| **Variables** | **95% CI for HR** | **p-value** | **c-index** |
| --- | --- | --- | --- |
| RV Global Strain | 1.18 (1.07-1.29) | 0.0005* | 0.77 |
| RV Free Wall Strain | 1.15 (1.06-1.23) | 0.0003* | 0.78 |
| Ejection Strain Rate | 1.05 (1.02-1.08) | 0.003* | 0.75 |
| Passive Filling Strain Rate | 0.98 (0.96-1.00) | 0.08 |  |
| Active Filling Strain Rate | 0.99 (0.96-1.01) | 0.2 |  |
| TAPSE | 0.96 (0.92 – 1.00) | 0.06 |  |
| TAPSE/sPAP | 0.1 (0.012–0.74) | 0.02 | 0.71 |
| RVEF | 0.94 (0.91 – 0.97) | 0.0002 | 0.77 |
| RV ESVI | 1.02 (1.01 – 1.02) | p<0.0001* | 0.82 |
| SVI | 0.96 (0.91 – 1.01) | 0.08 |  |
| RA pressure | 1.15 (1.08 – 1.22) | p<0.0001* | 0.76 |
| RV Global Strain/sPAP | 0.01 (0.0003-0.4) | 0.01 | 0.75 |
| SV/ESV | 0.06 (0.01-0.3) | 0.001 | 0.77 |
| Ees/Ea<sup>#</sup> | 0.70 (0.32-1.53) | 0.4 |  |
| Eed | 2.44 (1.44 – 4.14) | 0.0009 | 0.73 |
| **Strain Patterns** |  |  | 0.67 |
| Strain Pattern 1 (referent) |  |  |  |
| Strain Pattern 2 | 1.33 (0.22-7.964) | 0.7 |  |
| Strain Pattern 3 | 4.779 (1.06-21.6) | 0.042 |  |
| **FW Groups** |  |  | 0.78 |
| FW-G1 | (referent) |  |  |
| FW-G2 | 0.51 (0.05-5.65) | 0.9 |  |
| FW-G3 | 8.47 (1.91-37.6) | 0.005 |  |
| **Eed Groups** |  |  | 0.72 |
| Eed-G1 | (referent) |  |  |
| Eed-G2 | 1.57 (0.26-9.41) | 0.6 |  |
| Eed-G3 | 6.75 (1.49-30.5) | 0.01 |  |
| **FW strain and Eed** |  |  | 0.78 |
| RVFW>20 or Eed < 0.4 | (referent) |  |  |
| RVFW<20 or Eed > 0.4 | 1.2 (0.20-7.2) | 0.8 |  |
| RVFW<20 and Eed>0.4 | 14.0 (3.9 – 50.5) | <0.0001 |  |

**Figure 5.**
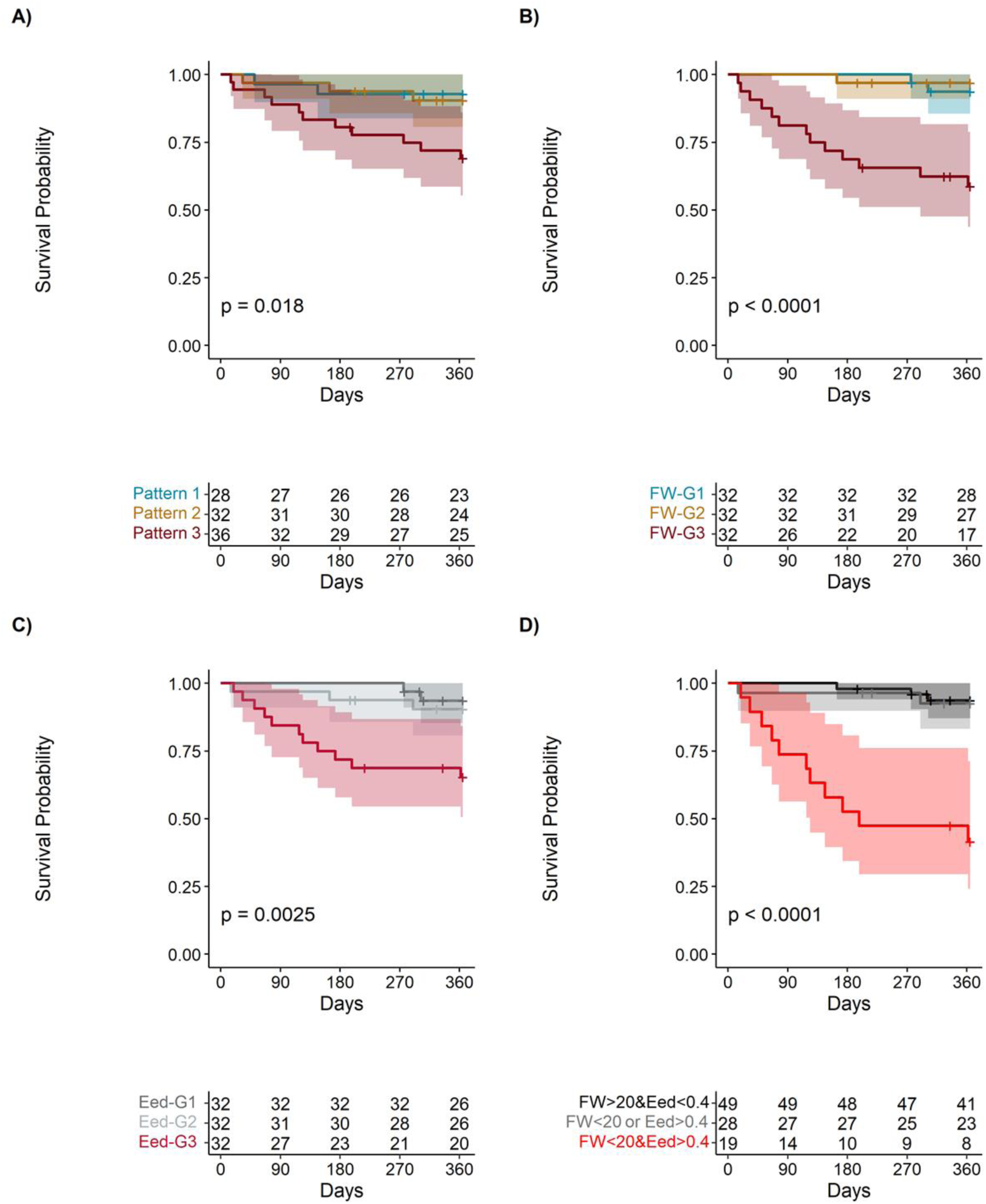
Associations of strain and diastolic stiffness with mortality. Strain Patterns (A), Free wall strain groups (B) and diastolic stiffness groups (C) significantly associate with one-year all-cause mortality. Strain Pattern 3, decreased free wall strain (FW-G3) and increased diastolic stiffness (Eed-G3) had decreased survival compared to the other groups. **D)** Participants with decreased free wall strain and increased diastolic stiffness (FW strain < 20% & Eed > 0.4 mmHg/ml: red) had significantly decreased survival compared to participants with FW strain >20% and Eed < 0.4 mmHg/ml (dark gray). Participants with only decreased strain (FW strain <20%) and low diastolic stiffness (Eed < 0.4 mmHg/ml) or only increased Eed (Eed > 0.4 mmHg/ml) and high FW strain (FW strain > 20%) did not have a decrease in survival at one-year.

## Discussion

We examined RV strain from four-chamber CMR images to assess phasic RV function across the cardiac cycle. Our results demonstrate that 1) RV global and free-wall strain derived from 4CH CMR images associates with RV ejection fraction and mortality, 2) Strain patterns as surrogates to describe phasic RV function by themselves are not enough to describe RV dysfunction, and 3) participants with reduced ventricular movement (RV FW strain <20% or RVEF <35%) and stiffer ventricles (Eed > 0.4) have worse one-year outcomes compared to other participants.

### Cardiac MRI assessments of RV strain

RV strain allows for the evaluation of the mechanical function of the RV.^20^ Because it normalizes movement to length/size, strain is also an angle independent measure. Assessment of RV strain from CMR imaging allows for better visualization of RV endocardial borders that is sometimes challenging with echocardiography especially in patients with a dilated RV or factors that lead to sub-optimal windows. Multiple methods are available for assessment of RV strain from CMR imaging including feature/tissue tracking in the cardiovascular imaging version 42 (cvi42, Circle Cardiovascular Imaging, Calgary, Alberta, Canada)^8^ or QStrain software (Medis BV, Leiden, The Netherlands).^21^ While full feature tracking or speckle tracking is the gold standard for assessment of strain, simplified methods to assess strain have been shown to have good reproducibility, shorter analysis time and comparable diagnostic and prognostic capabilities.^21,22^ The current analysis used RV endocardial length from the four-chamber CMR views in the calculations for RV global longitudinal strain.

Normal reference values for RV strain have been found to be both sex- and region-specific where the upper limits of normal for RV global strain were found to be -20% for men and -20.3% for women.^23^ The upper limits of normal for RV free wall strain was slightly higher at -22.5% for men and -23.3% for women.^23^ While there were no sex-associated differences in free wall strain in our cohort, there were significant changes in strain between the PH hemodynamic classifications (**Figure S5**). In our cohort, RV global strain decreased from 27.6 ± 4.5% in participants without PH (mPAP < 20 mmHg) to 19.7 ± 5.3% in the pre-capillary PH group and 18.3 ± 5.3% in the combined pre-post capillary PH group. This decrease in strain is reflective of the different ranges of identified prognostic cut-off values for RV strain depending on cardiovascular disease (RV free-wall cut-off for PAH: 12.5 to 25.2%, Heart Failure: 13.1 to 19%, and valvular disease: 15 to 23%).^20^

Regional changes in RV function were further investigated by breaking global strain into free wall and septal strain. RV free wall strain strongly correlated with global strain (r = 0.95, p<0.001) and varied from 7.3 to 42.9%. The cut-off point between FW groups 2 and 3 was 19.5% that is very similar to the RV free wall strain cutoff of 19% that has been found to significantly associate with mortality.^14^ Contrary to free wall strain, septal strain did not associate with global strain (r = 0.40, p<0.001). Septal strain was significantly decreased in FW-G3 (13.7 [11.5-16.3]%) compared to FW-G1 (18.1 [15.1-21.5]%) suggesting normal septal function until longitudinal systolic function is impaired. Longitudinal strain has been shown to significant correlate with RV hypertrophy (RV mass diastolic/BSA, rho = 0.63) and RV ejection fraction in patients with pulmonary hypertension.^8^ We found similar correlations between RV free wall strain and RV ejection fraction (rho = 0.8, p <0.0001) in our cohort. (**Figure S6**). When we used an RVEF cut-off of 35% instead of an RV FW strain of -20% in combination with diastolic stiffness (Eed > 0.4 mmHg/ml), there were only 3 of participants that had and RV free wall strain that met the cut-off of <20% but RVEF that did not (≥35%) (**Figure S4B**).

### Phasic Ventricular Strain to define RV adaptation

Strain analysis quantifies the contraction and relaxation of the RV and can be analyzed in in the longitudinal, circumferential, and radial directions.^24,25^ A shift from longitudinal to circumferential/radial movement is suggested to be a compensatory adaptation in age and increased afterload.^24–26^ In young, healthy individuals, RV movement is primarily in the longitudinal direction (base to apex) with some circumferential and radial movement.^24^ Normal age-associated reductions in RV longitudinal movement is associated with an increase in circumferential and radial movement.^24^ This shift from longitudinal to circumferential/radial movement has also been characterized in patients with transposition of great arteries and systemic right ventricles^26^ and patients undergoing transcatheter tricuspid valve repair ^25^. Diastolic dysfunction is associated with changes in the post-systolic filling patterns with impairments in the first phase of diastolic relaxation.^9,11^ Later stages of RV dysfunction with impairments in both systolic and diastolic function are associated with reduced global strain that is accompanied by impairments in both the passive and active diastolic filling (Strain Pattern 3). The later stages or the maladaptive RV phenotype is more clinically obvious with increased RV end-diastolic volume, reduced RV ejection fraction, elevated filling pressure and RV diastolic stiffness. The combined decrease in RV systolic and diastolic function translate to equal passive and active filling strain rates. ^9,24–26^ Similar strain patterns were found in patients undergoing transcatheter tricuspid valve repair.^25^ Patients with preserved longitudinal (TAPSE>17 mm) and global (RVEF >45%) function had preserved longitudinal, circumferential, and radial strain. However, in participants with preserved global function (RVEF > 45%) impairments in longitudinal function (TAPSE <17 mm) were compensated for with increased strain/movement in the radial and circumferential directions. When longitudinal and global function were both impaired, longitudinal, circumferential, and radial strain was reduced in all directions. The addition of strain pattern analysis have a potential added benefit when assessing patients that have increased pulmonary vascular resistance but preserved RV systolic and diastolic function.^9^ Strain patterns can further phenotype even with reduced free wall strain (**Figure S3**).

### Interactions between diastolic stiffness and strain

Due to the complex pathophysiology, RV dysfunction needs to be assessed by a combination of variables that incorporate systolic, diastolic and global function. Prognostic models that incorporate measures of systolic function (RV longitudinal strain, RVEF) and diastolic function (RA function, RA reservoir strain, tricuspid regurgitation and RA pressure) are more predictive of outcomes than hemodynamic variables and standard echocardiography variables alone.^9,11,14^ Specifically in Ghio et al. they found the combination model of advanced echocardiography variables was more predictive of outcomes(c-index:=0.726) compared to their models using hemodynamic variables(c-index=0.689) and standard echocardiography variables alone (c-index=0.667).^14^ Using combined criteria of decreased systolic function (RV free wall strain < 0.2) and increased diastolic stiffness (Eed > 0.4), we identified participants that had increased one-year mortality. There were no significant differences in one-year mortality in the other participants suggesting additional measures like strain patterns are needed in an RV with adaptive remodeling.^9^ Additionally, one-year mortality was not significant across PH types (**Figure S5**). Individual components of the strain patterns including the passive and active filling rates did not individually associate with outcomes (**Table 3**). Looking at the RV function variables that had been included in the risk assessment in the 2022/2023 pulmonary hypertension guidelines, there is discordance in how participants would be classified into low, intermediate and high risk across the variables (**Figure S4**). Additional work is needed to develop better models to classify risk when there is contradictory findings across the RV function measures.

### Limitations

This was a small, retrospective, and cross-sectional cohort study in participant across the WSPH classification spectrum. In the survival analysis, survival bias may be present, as follow-up time did not include the time from the initial diagnosis to catheterization and imaging. The combination of RHC and CMR imaging information allows us to better describe the complexity of diastolic function but the small sample size limits the broad application of the findings. RV strain is usually assessed using speckle tracking echocardiography but that analysis requires extra training and experience to get reproducible results that typically limits the analysis to specialized centers. Assessment of RV strain from CMR 4CH cine views has been shown to associate with more standard strain measures and can be accessible from standard cardiac MRI studies.^21,22^ CMR tagging allows for quantification of intramyocardial motion including strain encoding (SENC) but requires additional scans to be made.^27^ Another limitation in our analysis was that strain was only assessed in the longitudinal direction. More complete description of the pathological shift of phasic RV function will require the incorporations of circumferential and radial strain. More standardized methods for strain pattern determination will facilitate the reproducibility and adjudication between patterns. To minimize subjectivity, we used average strain rate during systole, early/late diastolic filling when assigning strain patterns. Replication of these methods in a larger multi-institution cohort will be necessary to validate the results.

### Clinical Relevance

Multimodal approaches improve the characterizing RV adaptation and dysfunction in patients with PAH. In the current cohort, RV strain associated with one-year mortality (P<0.001) where TAPSE did not (p = 0.06) potentially due to the normalization of movement in the ventricle relative to heart size. Free wall strain has been shown to associate strongly with mortality^12,13,28^ but has been shown to be afterload-dependent.^8,29^ Normalizing systolic RV movement (TAPSE or strain) for afterload (PA systolic pressure) provide simplified coupling parameters (TAPSE/PASP, RVGLS/PASP) that associate with mortality and glucose uptake in PET imaging.^30^ Patients with increased RV diastolic stiffness also have impaired RV filling (strain pattern 3) that when combined with decreased systolic function can lead to increased vena cava backflow and reductions in diastolic blood supply.^31^

### Conclusions

This study demonstrates that the multi-faceted approach of incorporating hemodynamics alongside imaging and strain parameters is important in patients with pulmonary hypertension. Assessment of phasic changes in ventricular function using ejection, passive filling and active filling strain/strain rates do provide additional pathophysiological information but assessment of strain patterns in patients with pulmonary hypertension are not sufficient for identifying reduced function. Deep phenotyping using a combination of RV strain and diastolic stiffness is highly selective of participants with increased one-year mortality.

## Data Availability

The data that support the findings of this study are available from the corresponding author upon reasonable request.

## Abbreviations

CO: Cardiac Output
CMR: Cardiac Magnetic Resonance
Ea: arterial elastance
Ees: End-systolic Elastance
Eed: end-diastolic elastance
ESP: end-systolic pressure
FAC: fractional area change
FW: Free wall
LD: Length during diastole
L0: Original length
PA: Pulmonary artery
PAWP: Pulmonary Artery Wedge Pressure
Pmax: Maximum isovolumetric pressure
PVRi: pulmonary vascular resistance index
Sep: Septum
TAPSE: Tricuspid Annular Plane Systolic Excursion

## Acknowledgements

We thank Orlando Simonetti, PhD and Juliet Varghese, PhD at The Ohio State University for their assistance with the OSU CMR registry.

## Supplemental Material

Tables S1–S4

Figure S1-S6

